## Supplementary material for "Comparison of stochastic and deterministic models for *gambiense* sleeping sickness at different spatial scales: A health area analysis in the DRC": SI text 1

**S1.2 Using the models** **6**

### S1.1 Data

#### S1.1.1 Formatting the data

For the present study we converted available data from the HAT Atlas [5, 8] into a format suitable for model fitting at both health zone and health area levels. This methodology is similar to the process described in Crump et al. [3], but using updated shape files, including health area boundaries, and an updated data set that includes reported cases and active screening data from 2000–2020.

The HAT Atlas data for the DRC from 2000–2020 were provided in a spreadsheet format. Records were gHAT case records, aggregated by year, surveillance type and location as defined by multiple fields.

We use shape files from American Red Cross (ARC) [2]. There are shape files containing boundaries of health zones (an organisational unit with a typical population size around 150,000) across the whole DRC, and health areas (sub-units of health zones, which cover the home location of approximately 10,000 people) for a large portion of the DRC. The health zone of interest, Mosango, was fully covered by these files.

Geolocation data were used where present to allocate data to the health areas in the shape file. If this information was not available for the record then other data (e.g. an OCHA file of geolocations of localities; and a file of geolocations of health facilities from the Global Healthsite Mapping Project [7]) were used to assist in matching and locating the gHAT data, by providing alternative spellings of names and potentially geolocations for non-geolocated gHAT locations. This process is very similar to that described previously [3].

#### S1.1.2 Health areas of Mosango health zone

In our presented analysis, we focus on Mosango health zone, which is comprised of sixteen health areas. In the main manuscript, and in particular in Fig 5, we label these sixteen health areas A1–A16 for brevity. The full names of these health areas are as follows:

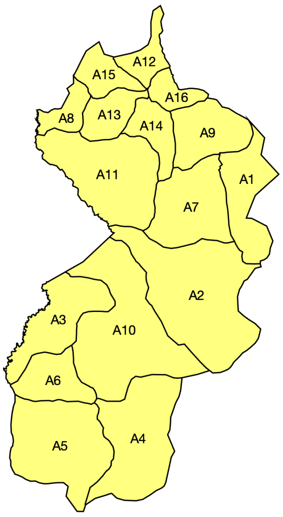

- A1 – Kasay
- A2 – Kinzamba I
- A3 – Kinzamba II
- A4 – Kinzanda
- A5 – Kinzenzengo
- A6 – Kipembe
- A7 – Kipwanga
- A8 – Kitambo
- A9 – Kumbi Mbwana
- A10 – Mangungu
- A11 – Mosango
- A12 – Mudiambu
- A13 – Muluma
- A14 – Muwanda Koso
- A15 – Tumikia
- A16 – Yenzi

Fig A: **Health area map for Mosango health zone.** Names of health areas in Mosango matched to labels given in main text. Shape files used to produce these maps were provided by Nicole Hoff and Cyrus Sinai under a CC-BY licence (current versions can be found at `https://data.humdata.org/dataset/drc-health-data`).

#### S1.1.3 Comparing health zone and health area data for Mosango

In comparing between health zone and health area data sets there are minor discrepancies in the values for various reasons. All these differences will not substantially affect the results, but are noted here.

The population sizes for health areas and health zones come from different sources. We have estimates based on 2013 data for health areas and 2015 data for health zones. Most of the difference in values between aggregated health areas and the health zone number is therefore accounted for in the assumption of 3% annual population growth, which reconciles the difference in year. However, there are some small difference, with the health zone estimate 2.9% larger than the aggregated health areas (Fig B).

Additionally, for the number of people actively screened, active cases, and passive cases, all data comes from the same source — the HAT Atlas [6, 8] — however, we could not assign all data to particular health areas. There are cases where a health zone is listed, but we an unable to determine the health area, within the health zone, the data came from. Therefore, figures for these data for aggregated health areas are slightly slower. In total there are 4555 (0.9%) of people actively screened, one (0.2%) active case, and six (0.8%) passive cases that could not be mapped to a health area (Fig B).

#### S1.1.4 Health areas with limited data

In Crump et al. [3], model fitting was not carried out on health zones that were determined to have insufficient data. The threshold was set at a minimum of ten data points to perform the analysis, where a data point consisted as a year where an active screening occurred or a year in which at least one passive case was detected. For the health zone of Mosango, this threshold was exceeded, but for the health areas in Mosango, not all meet this criteria. The health areas of Kasay and Kipwanga only have seven and nine data points respectively (Fig C).

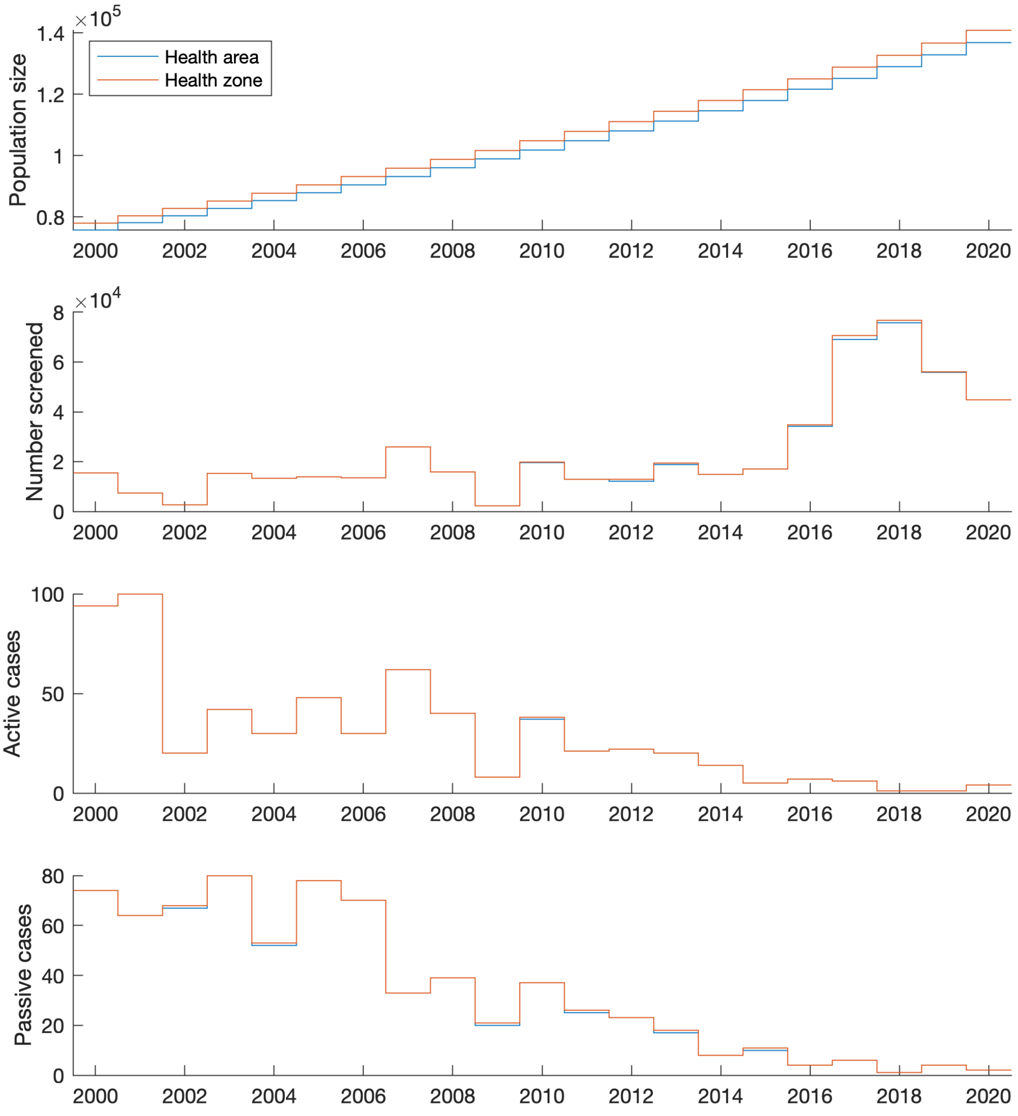

Fig B: **Data for Mosango health zone.** Data on population size, number of people actively screened, active and passive cases for Mosango health zone. The blue line shows aggregated health area data and the orange line shows health zone data.

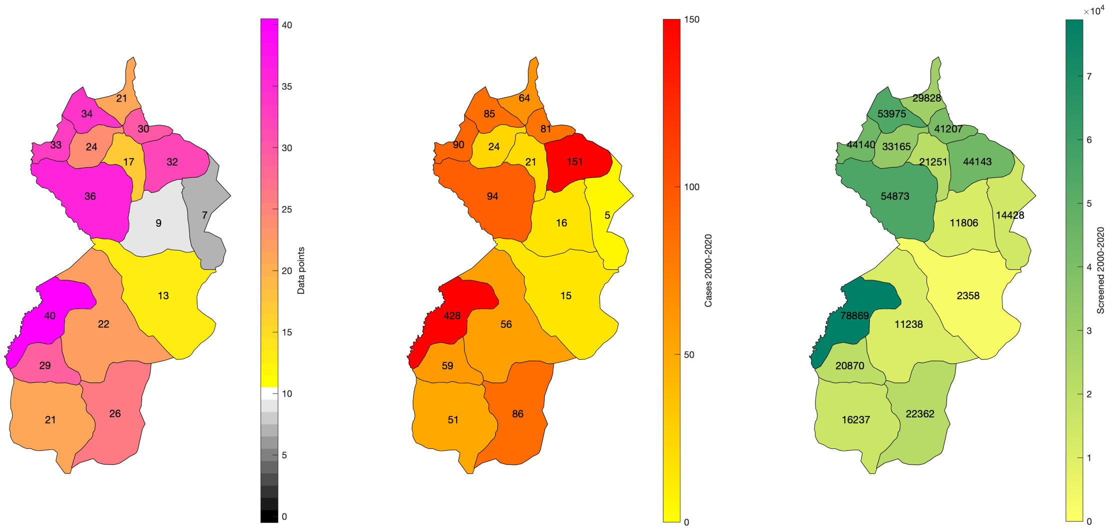

Fig C: **Maps of data points in Mosango health zone.** The total number of data points, cases 2000–2020 and people actively screened 2000–2020 for all health areas of Mosango. Shape files used to produce these maps were provided by Nicole Hoff and Cyrus Sinai under a CC-BY licence (current versions can be found at `https://data.humdata.org/dataset/drc-health-data`).

In the main manuscript of this paper, we determine all health areas to have sufficient data and so choose to fit the model to these data in the same way as any other health area. However, to perform extra analysis about this criterion, we additionally fit a model to the merged data from Kasay and Kipwanga, since they share a boundary and together contain 13 data points (Fig D). The results show that both models (either aggregated or fitted to merged data) provide very similar outputs, with almost completely overlapping box plots, particularly as the numbers are so small. This provides compelling evidence that these separate health area fits provide good results, despite the small data sets in these locations.

#### S1.1.5 Time step of stochastic models

Previous stochastic modelling work [1, 4] has used a time step of one day for simulating the tau-leaping algorithm. To save computation time, with the more intensive stochastic fitting process, we change this to a five day time step. This is still sufficiently small as to remain a good approximation of the full Gillespie algorithm, due to the long time scales of gHAT infection.

We show that there is negligible difference between the number of active and passive cases detected, as well as the probability of EoT, when stochastic projections are simulated using time steps of $\tau = 1$ day and $\tau = 5$ days (Fig E). The active case distributions are identical and there is a maximum discrepancy in one passive case in the median values based on 100 stochastic simulations for 2,000 parameter sets sampled from the posterior values. Also, for each year the maximum absolute difference in PEoT values is $0.005$, and hence the curves look indistinguishable on the right panel of Fig E. Therefore, we believe these differences are sufficiently small to be accounted for by the stochastic noise and so hence have used the 5 day time step in all simulations in this manuscript.

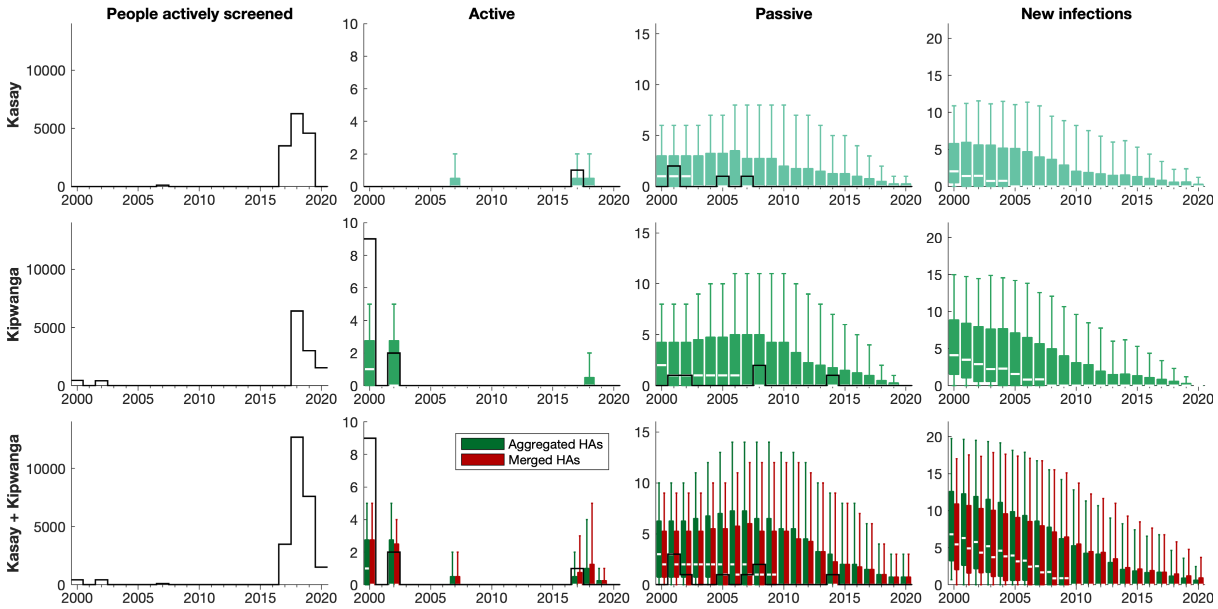

Fig D: **Kasay and Kipwanga health areas.** Annual people actively screened, active and passive cases and new infections for Kasay, Kipwanga and both Kasay and Kipwanga health areas. Aggregated results in the bottom row show the sum of the first two rows, while merged results show new fitting for the data from both health areas.

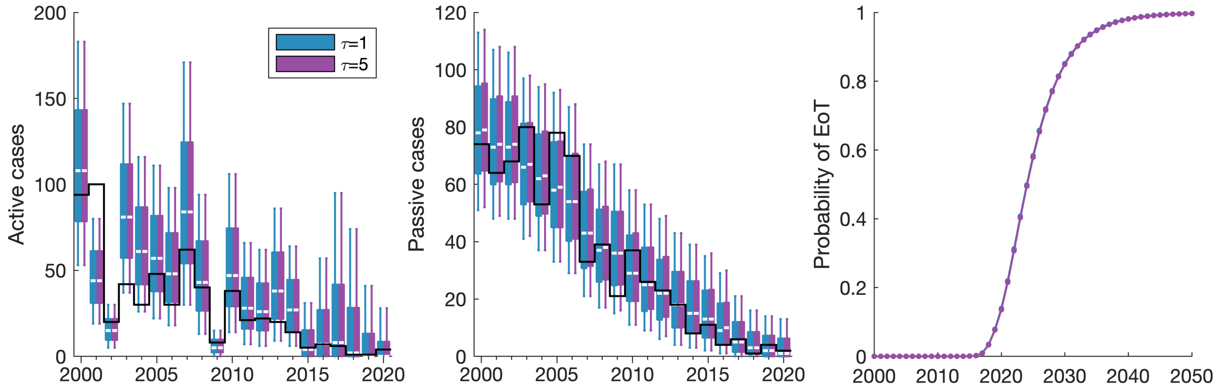

Fig E: **Difference in using $\tau$-leaping time step of one or five days for stochastic simulations.** Outputs show the distribution of the number of active and passive cases for both time steps, along with curves for the probability of EoT.

### S1.2 Using the models

All models presented were simulated assuming that, prior to 1998, they were at their endemic equilibrium. This endemic equilibrium was calculated analytically, based on model parameterisation, and used as an initial condition in the model code. In 1998 we assume active screening began at the same level reported in 2000 and that there was improvement to passive screening due to the availability of the CATT diagnostic test. In our model this has the effect of perturbing the dynamics away from endemic equilibrium and reducing transmission.

For fitting the models, there are two elements, each of which is initialised.

- Two chains are run in the Markov chain Monte Carlo (MCMC) used in the fitting. The chains are initialised using the fixed parameters and by random perturbations around supplied, individually valid, initial values of each parameter being fitted, rejecting those parameter sets that do not produce a valid posterior probability.
- Each projection is initialised with a randomly sampled realisation from the posterior distribution of fitted parameters alongside the set of fixed parameters.
