## Supplementary material for "Comparison of stochastic and deterministic models for *gambiense* sleeping sickness at different spatial scales: A health area analysis in the DRC": SI text 2

#### Contents

**S2.1 Comparison of model variants** 1

**S2.2 Example health area projections** 5

**S2.3 All health area model outputs** 7

#### S2.1 Comparison of model variants

A direct comparison of the posterior outputs of the deterministic and stochastic fitting shows very similar results between the two approaches in the Mosango health zone (Fig A). There is a slight shift to smaller values of $\gamma_{H_{\text{amp}}}$, which denotes the relative improvement in the passive stage 2 detection rate, but otherwise the posteriors are almost identical.

In addition to the outputs shown in Fig 6 and Fig 7 of the main manuscript, we perform both deterministic and stochastic model fitting on both health area and health zone data, as well as using the deterministic model fitting output posteriors in the stochastic model. This determines whether there is a large impact in fitting directly to a deterministic or stochastic model, or whether using the same parameters in each model variant could be sufficient.

In both health zones (Fig B) and aggregated health areas (Fig C), we see that using parameters from the deterministic fitting in the stochastic model variant provides much more similar outputs to the direct stochastic model fitting than the direct deterministic fitting. This indicates that most of the difference in model output comes from the choice of deterministic or stochastic, rather than the parameters used (when a model variant has been fitted to data). It is particularly notable that there is a large increase in uncertainty under stochastic projections.

The probability of EoT increases from zero to one much faster under the deterministic variant (evidenced by the smaller uncertainty in the box plots), whereas both methods that use stochastic projections show very similar results.

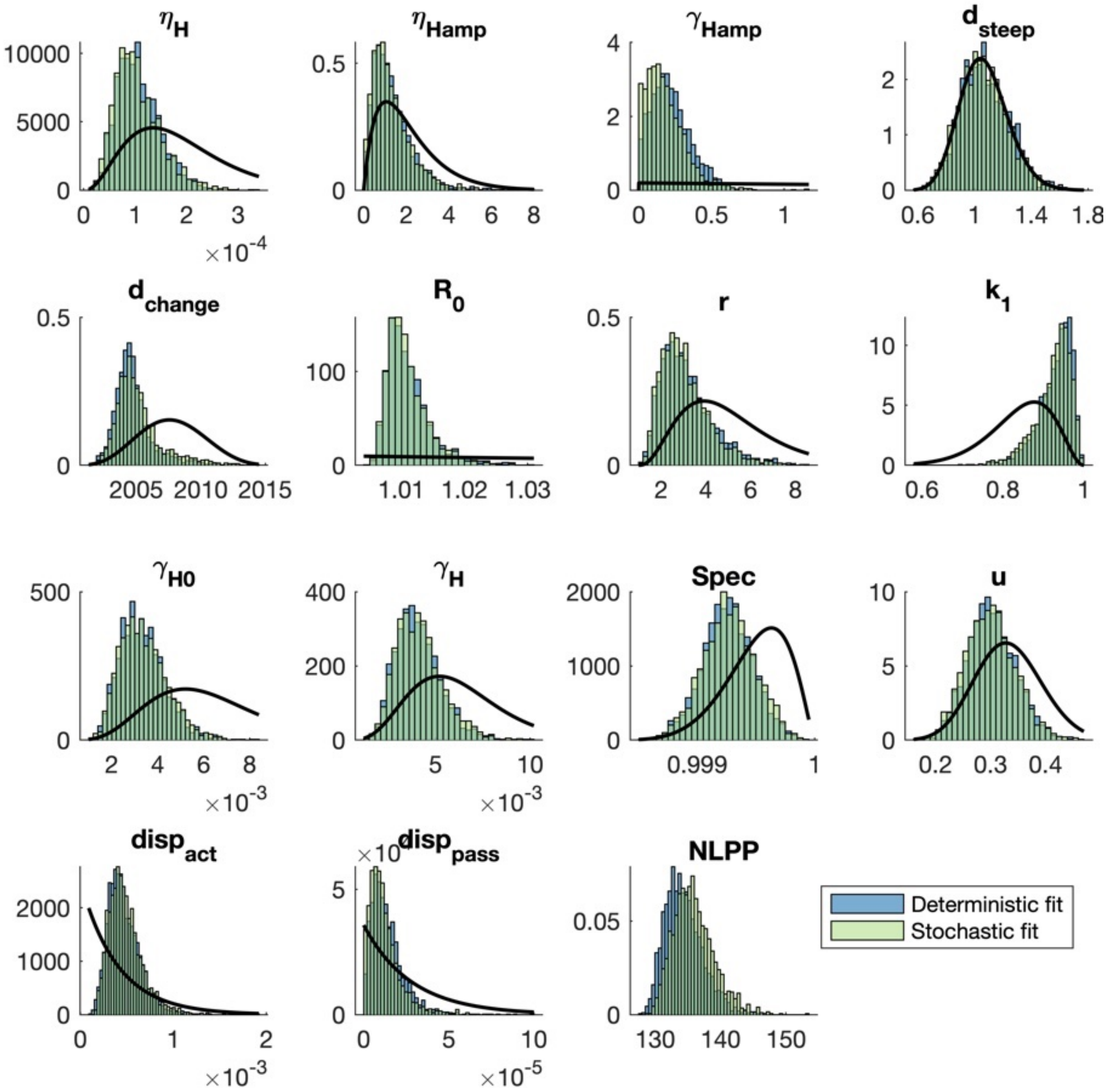


Fig A: **Difference between deterministic and stochastic posteriors for Mosango health zone.** The posterior distribution of all 14 fitted parameters are shown along with the negative log posterior probability (NLPP). The solid black lines show the prior distributions.

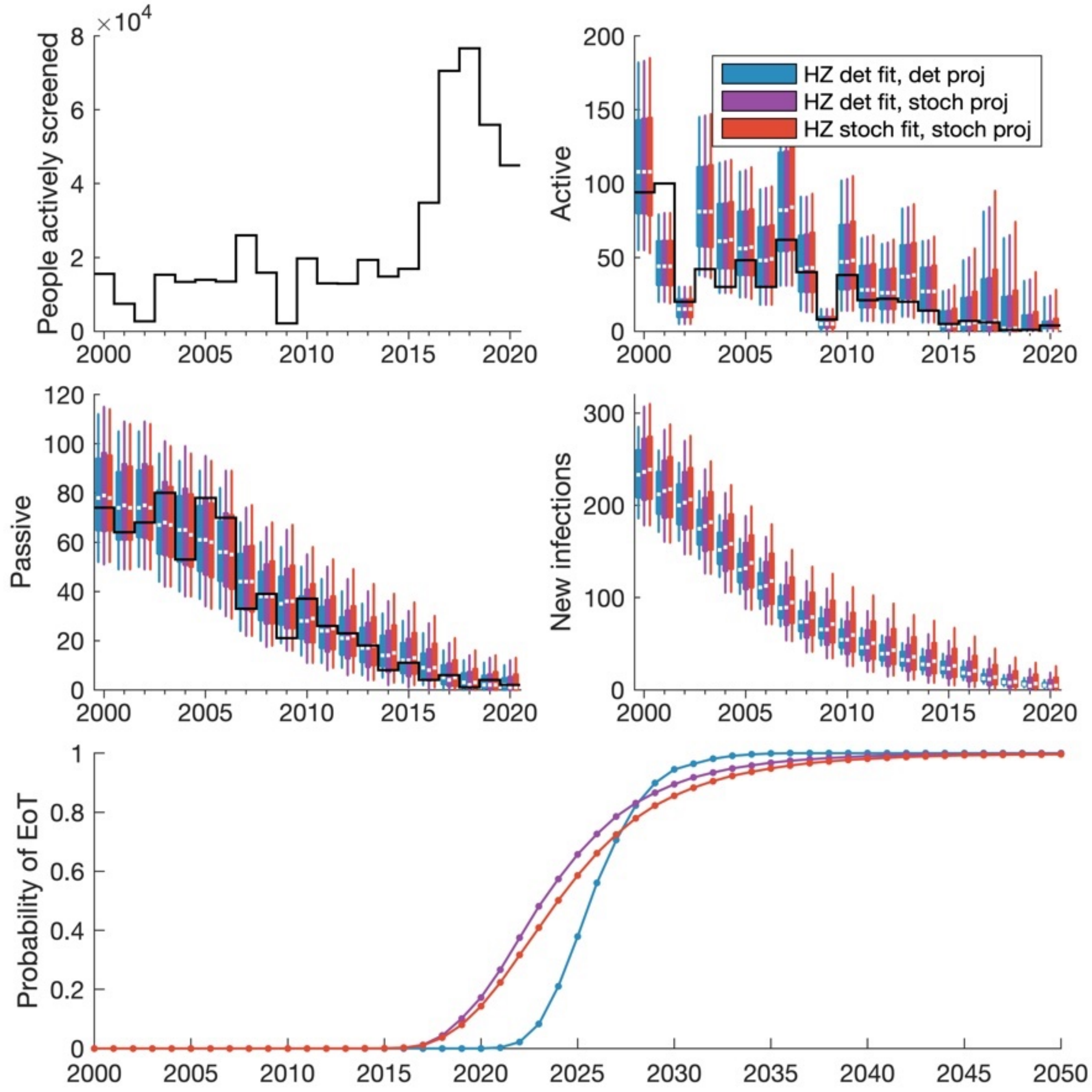


Fig B: **Different combinations of fitting and projection methods for the health zone level.** The coloured box plots and line plots show annual model outputs for different fitting approaches, where blue boxes show the deterministic model, purple shows the deterministic model posteriors in the stochastic model and red shows the stochastic model. Data for the number of people actively screened and the active and passive cases reported is displayed as solid black lines. The median value of the box plots is shown as a white line, with box plot whiskers representing 95% credible intervals.

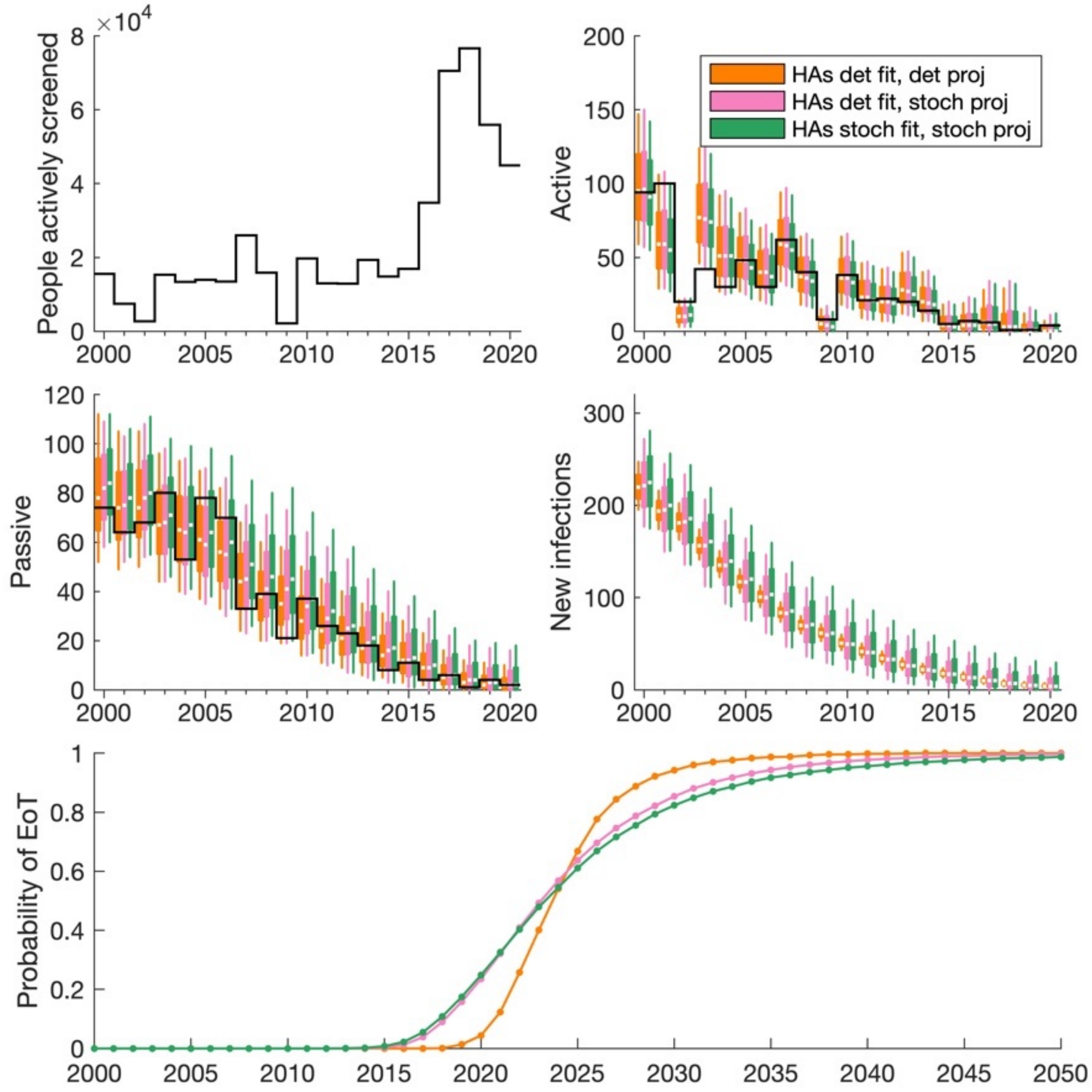


Fig C: **Different combinations of fitting and projection methods for the health area level then aggregating for the health zone.** The coloured box plots and line plots show annual model outputs for different fitting approaches, where orange boxes show the deterministic model, pink shows the deterministic model posteriors in the stochastic model and green shows the stochastic model. Data for the number of people actively screened and the active and passive cases reported is displayed as solid black lines. The median value of the box plots is shown as a white line, with box plot whiskers representing 95% credible intervals.

#### S2.2 Example health area projections

One of the main benefits of being able to simulate infection dynamics that is mentioned in the main manuscript is the ability to have specific and different intervention strategies in each health area. The previous approach with health zones made the assumption that the intervention was uniform across the whole area. This is clearly not reality as additional active screening could be targeted towards certain communities, there is a variation in access to health facilities, and tsetse control will typically only be implemented along rivers.

Here we show the benefit of changing the interventions across health areas by varying the coverage of active screening (Fig D). We consider the future strategy, from 2023 onwards, of continuing active screening in Mosango with 48,000 people screened annually, but distributing this differently. In Fig D, the purple boxes show an equal distribution and hence 3,000 people screened in each location. However the blue and pink boxes show 5,000 people screened in eight health areas and 1,000 in the other eight health areas. In the blue boxes, these eight health areas where only 1,000 people are screened are chosen to be the top eight highest prevalence health areas for the pink boxes, with 5,000 people screened in the lowest prevalence health areas. The opposite is true for the pink boxes.

By targeting the high prevalence health areas, more cases are detected in active screening initially, which drives down the prevalence and limits onward transmission and hence decreases the number of new infections. In the long run, more cases have been treated and so fewer cases are detected in active screening at later time points. Conversely, targeting the low prevalence health areas results in fewer detected cases and so continued transmission, which results in more new infections. This remains the case throughout and, as such, there are more detected cases in later time points because elimination is less likely to have occurred. The EoT plot shows that the highest probability of EoT across each year is obtained with the strategy of targeting high prevalence health areas, whereas the lowest probability of EoT is under the strategy targeting low prevalence health zones.

While in this example, there is relatively low prevalence across the health zone and so elimination is likely anyway, the example highlights the gains that can be made by carrying out screening in the most impactful places and so demonstrates the need for these fine scale models.

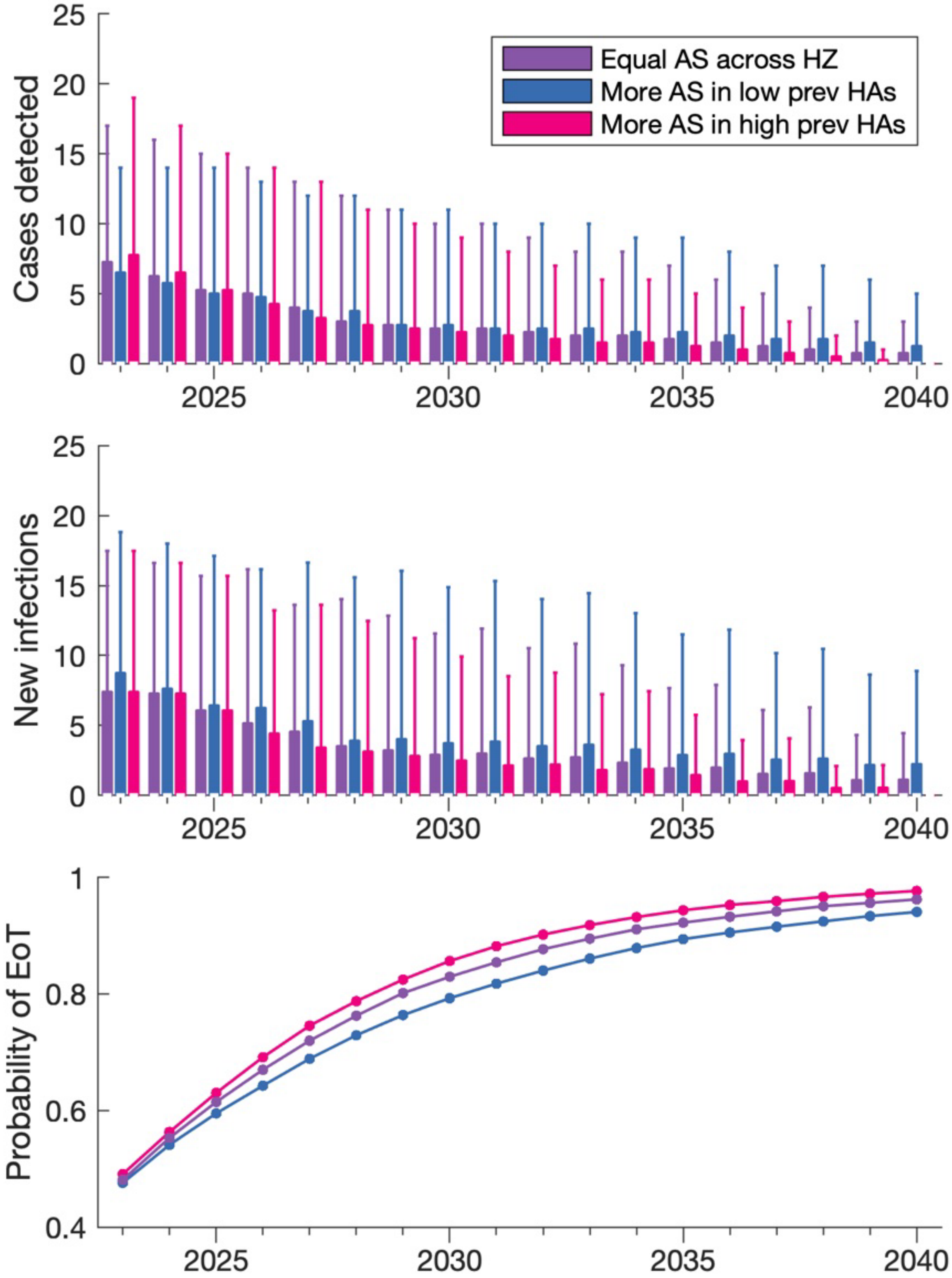


Fig D: **Future projections for varying active screening coverage in Mosango 2023–2040.** Box plots show the total number of cases detected (active and passive) and the number of new infections under different active screening strategies, with the probability of EoT shown in the bottom panel. Purple shows equal active screening coverage in all health zones, blue for where low prevalence health areas are targeted, and pink for targeting high prevalence health areas.

### S2.3 All health area model outputs

Figs E–T show the joint posterior from the pMCMC for each of the 16 health areas of Mosango along with the fit to active and passive case data.

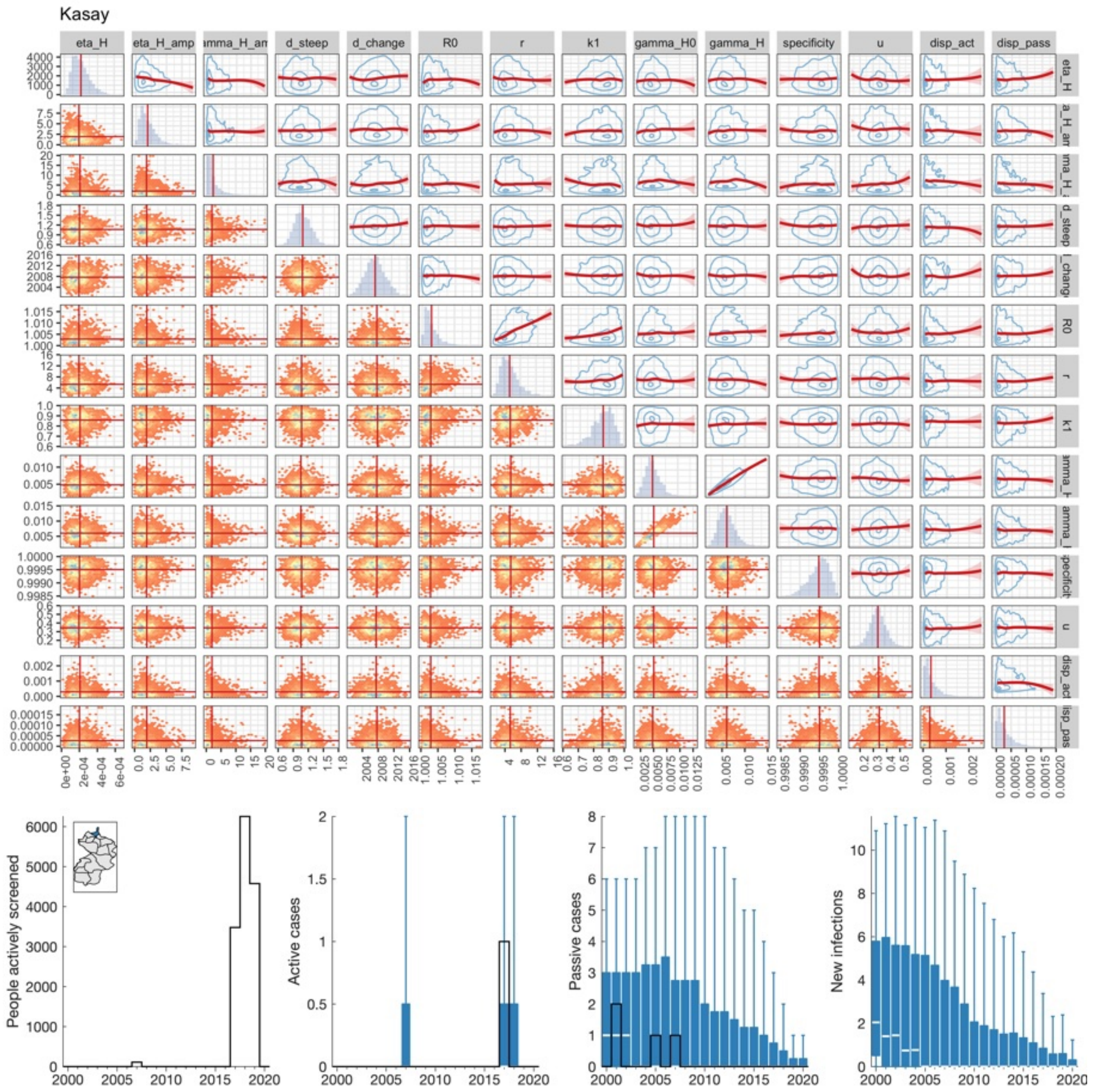


Fig E: **Kasay (A1) joint posterior and fit to data.**

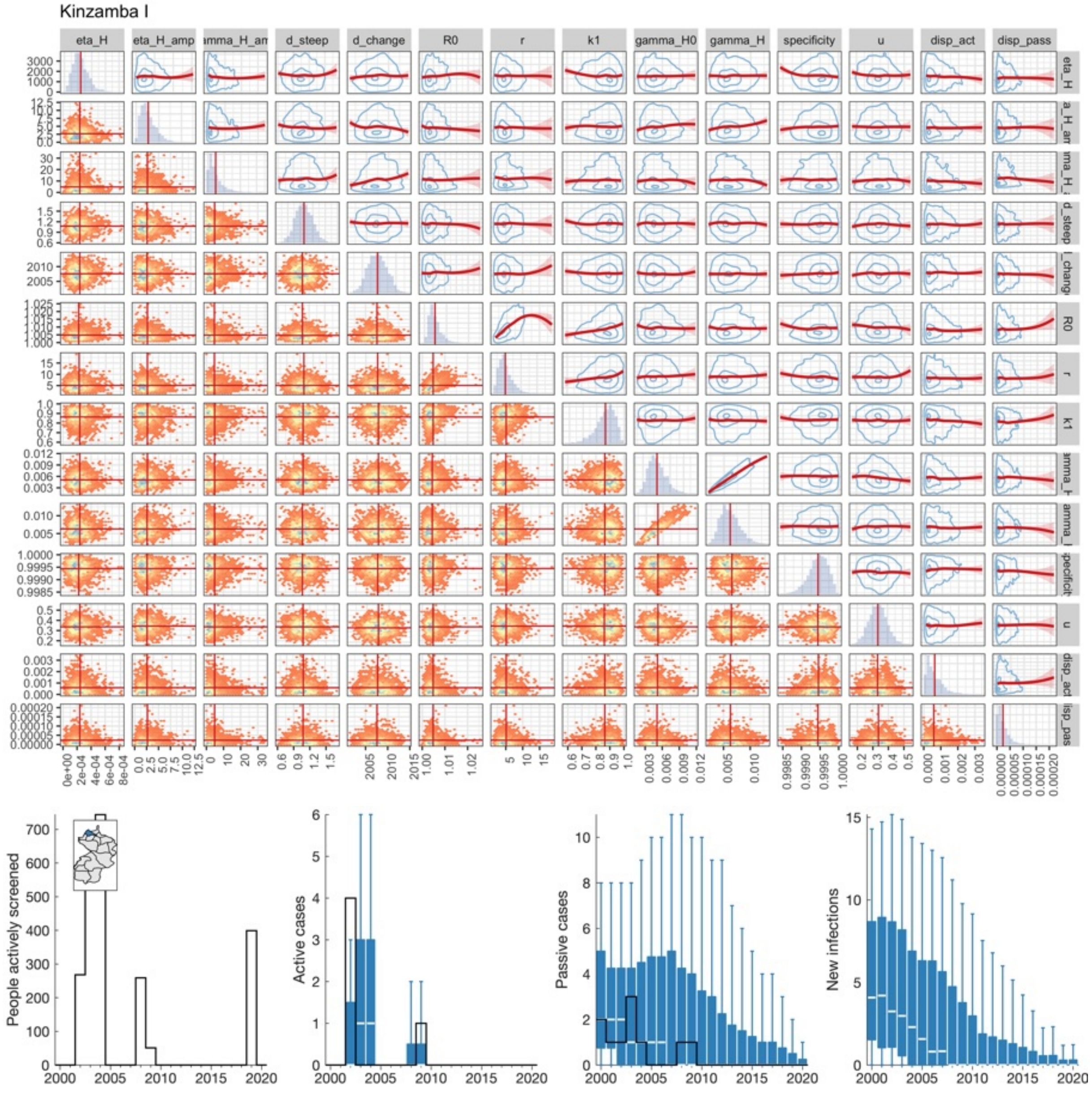


Fig F: **Kinzamba I (A2) joint posterior and fit to data.**

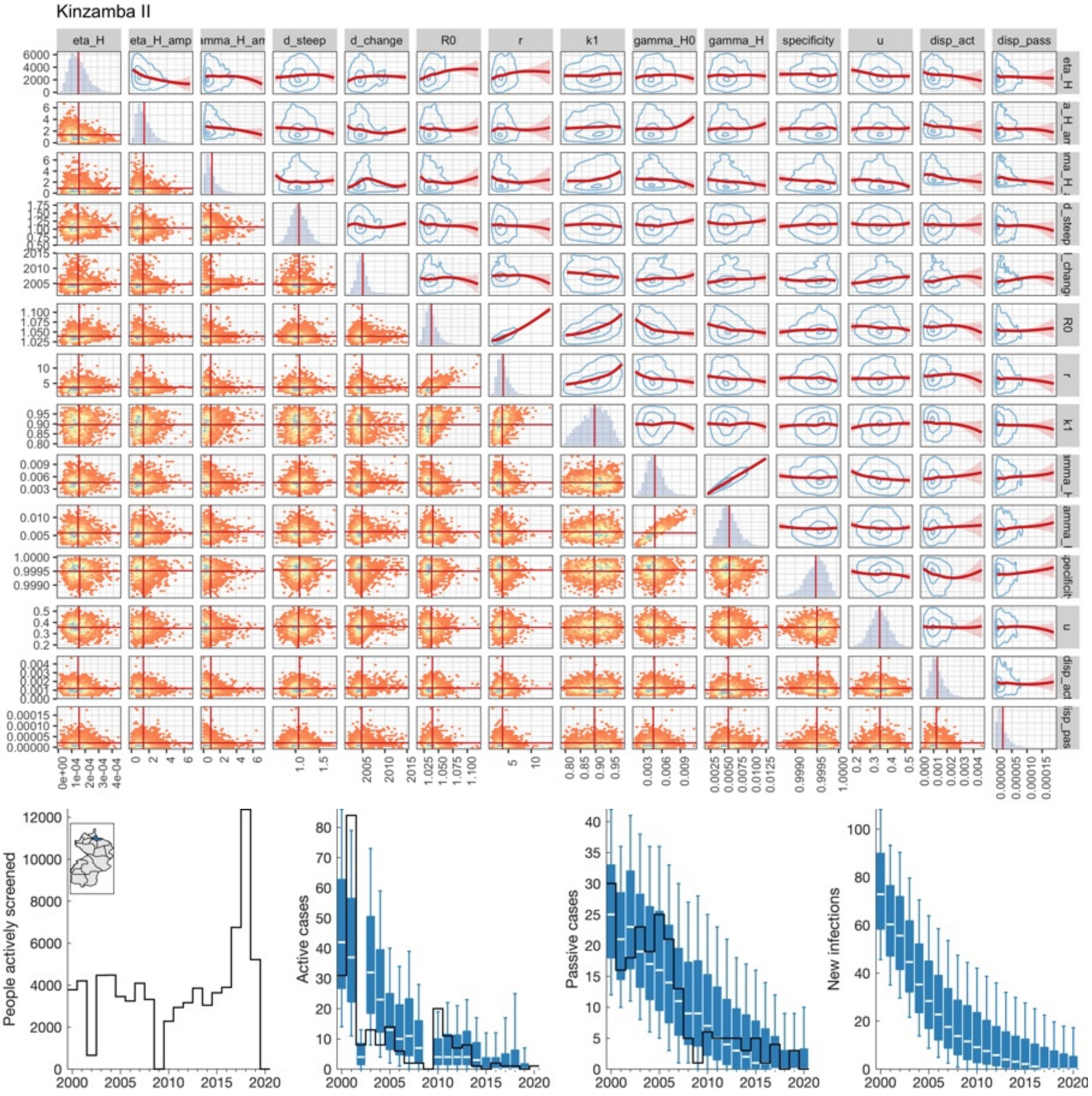


Fig G: **Kinzamba II (A3) joint posterior and fit to data.**

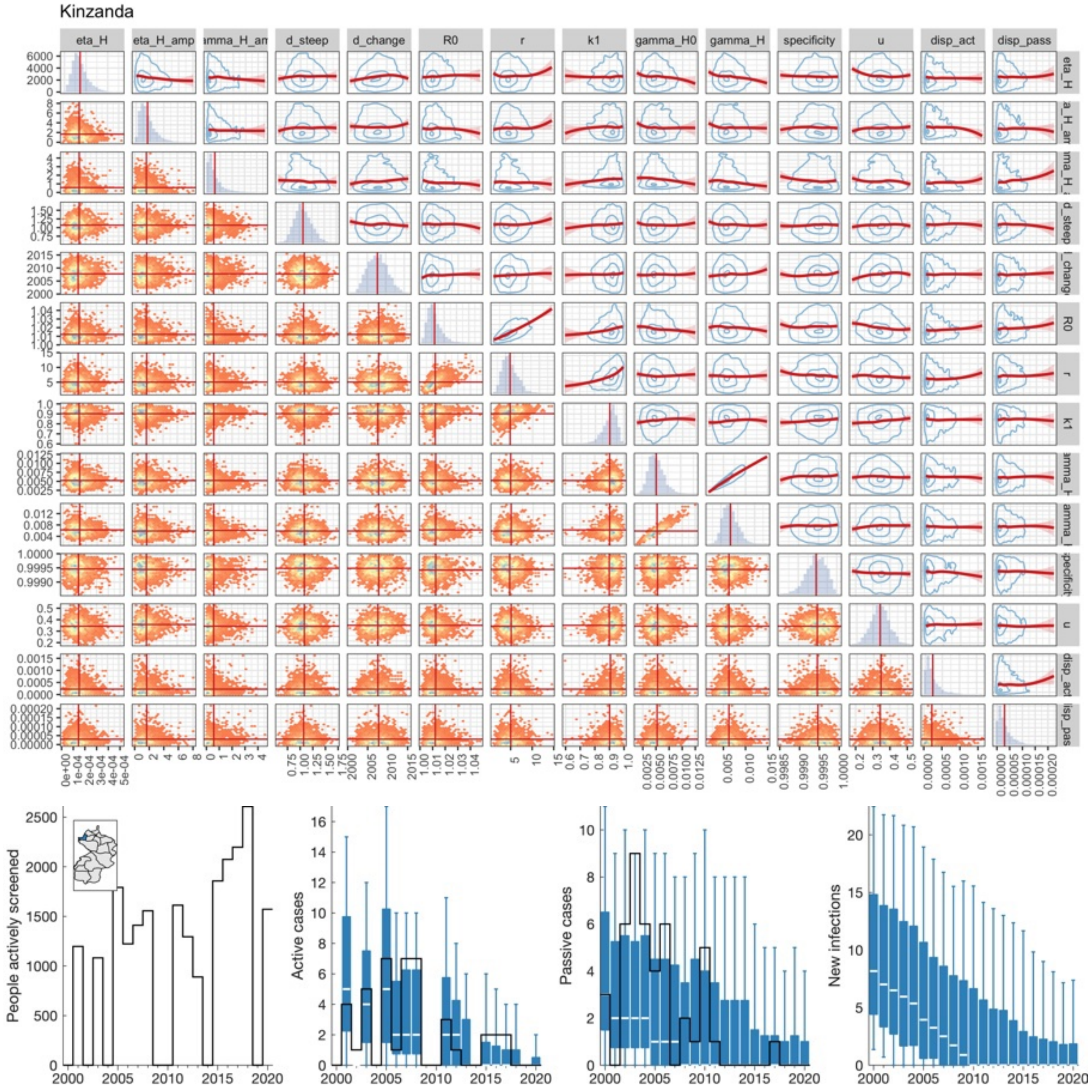


Fig H: **Kinzanda (A4) joint posterior and fit to data.**

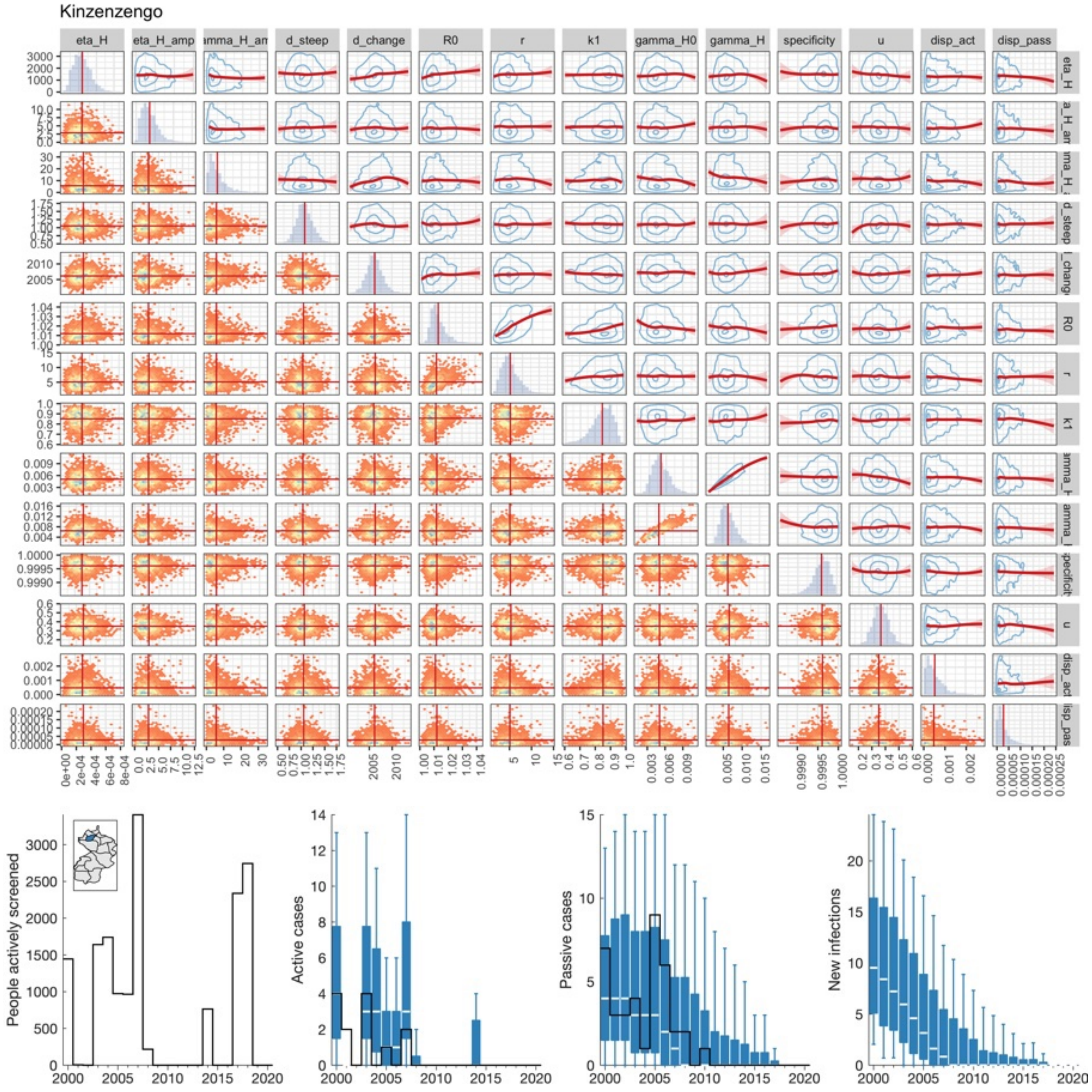


Fig I: **Kinzenzengo (A5) joint posterior and fit to data.**

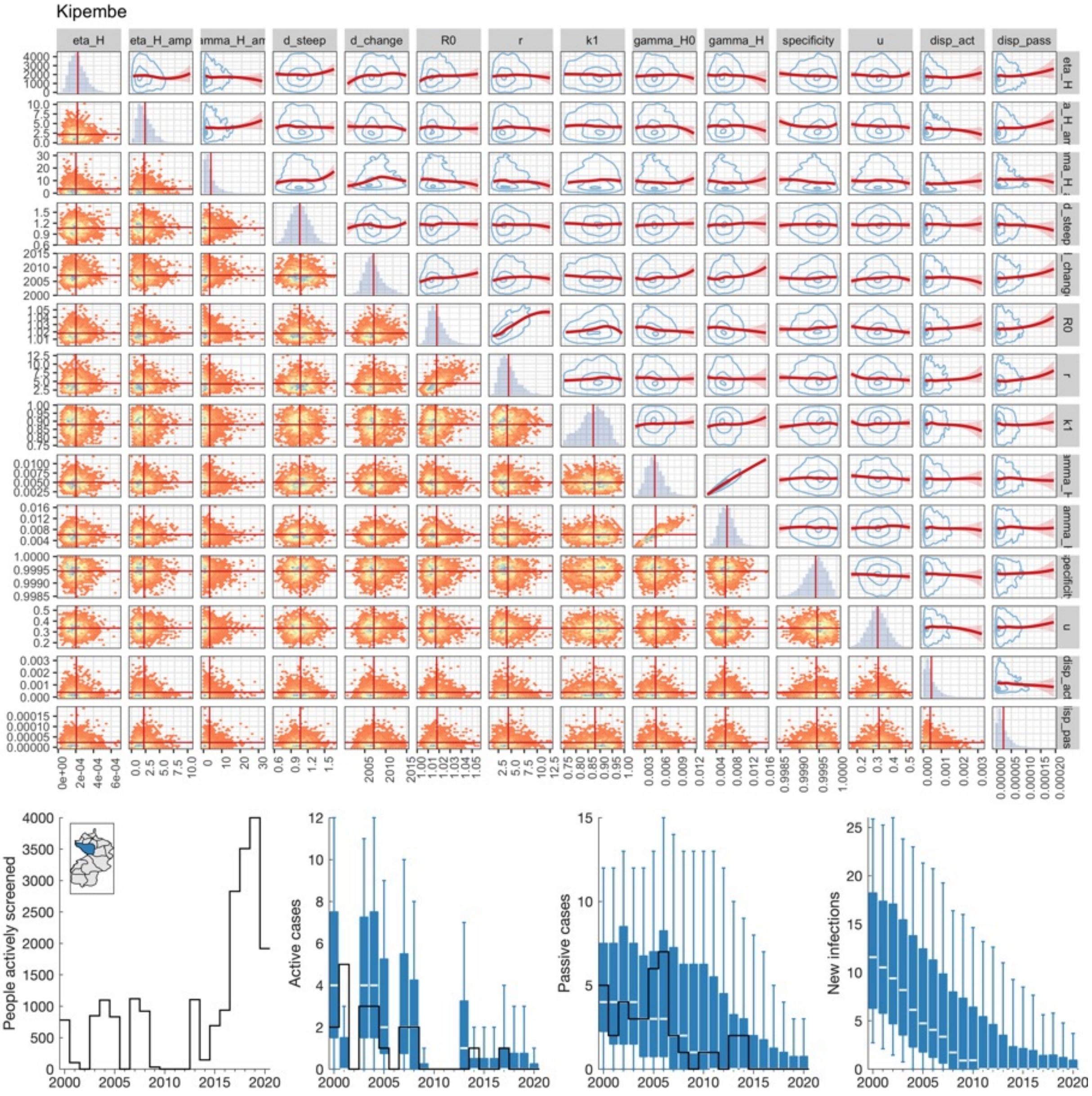


Fig J: **Kipembe (A6) joint posterior and fit to data.**

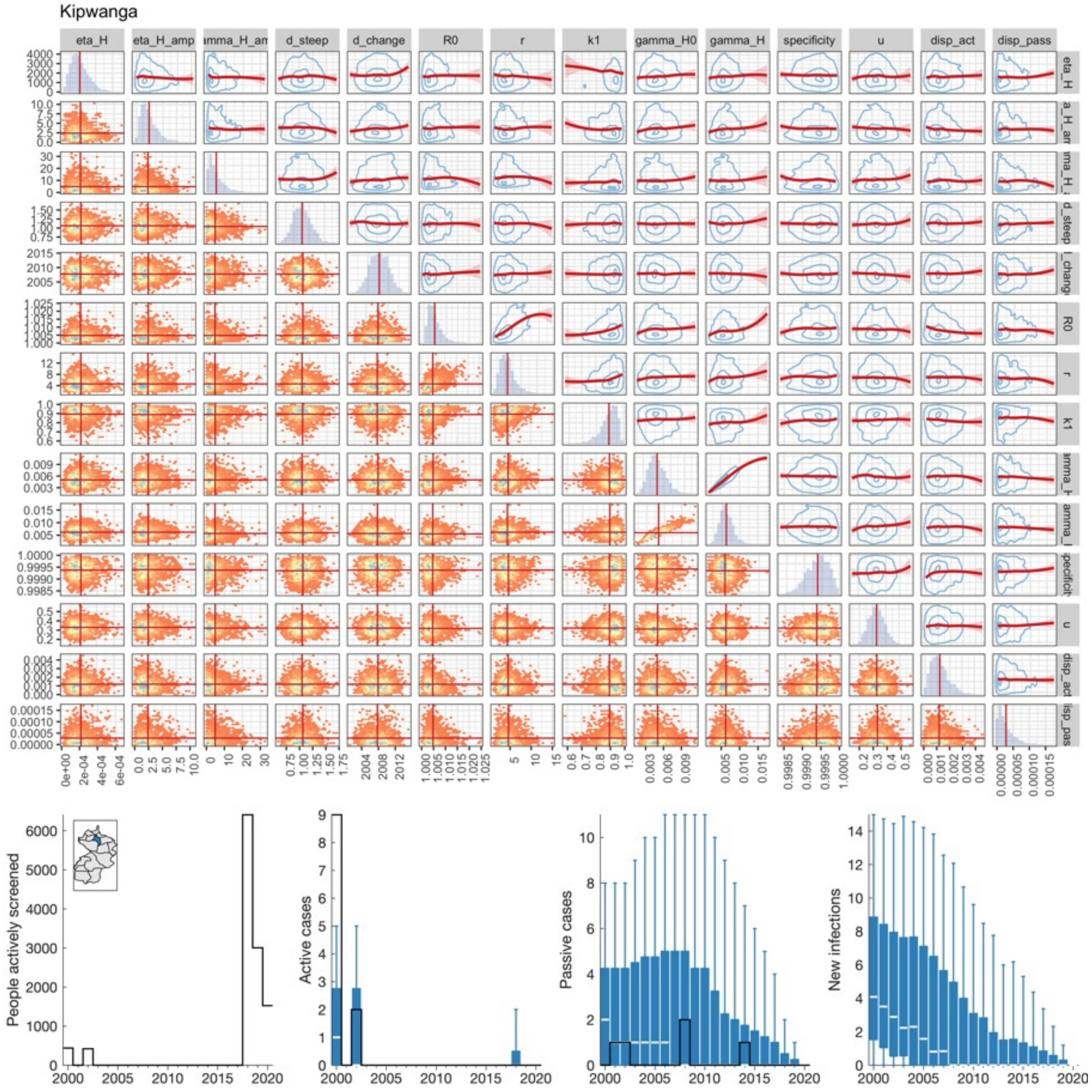


Fig K: **Kipwanga (A7) joint posterior and fit to data.**

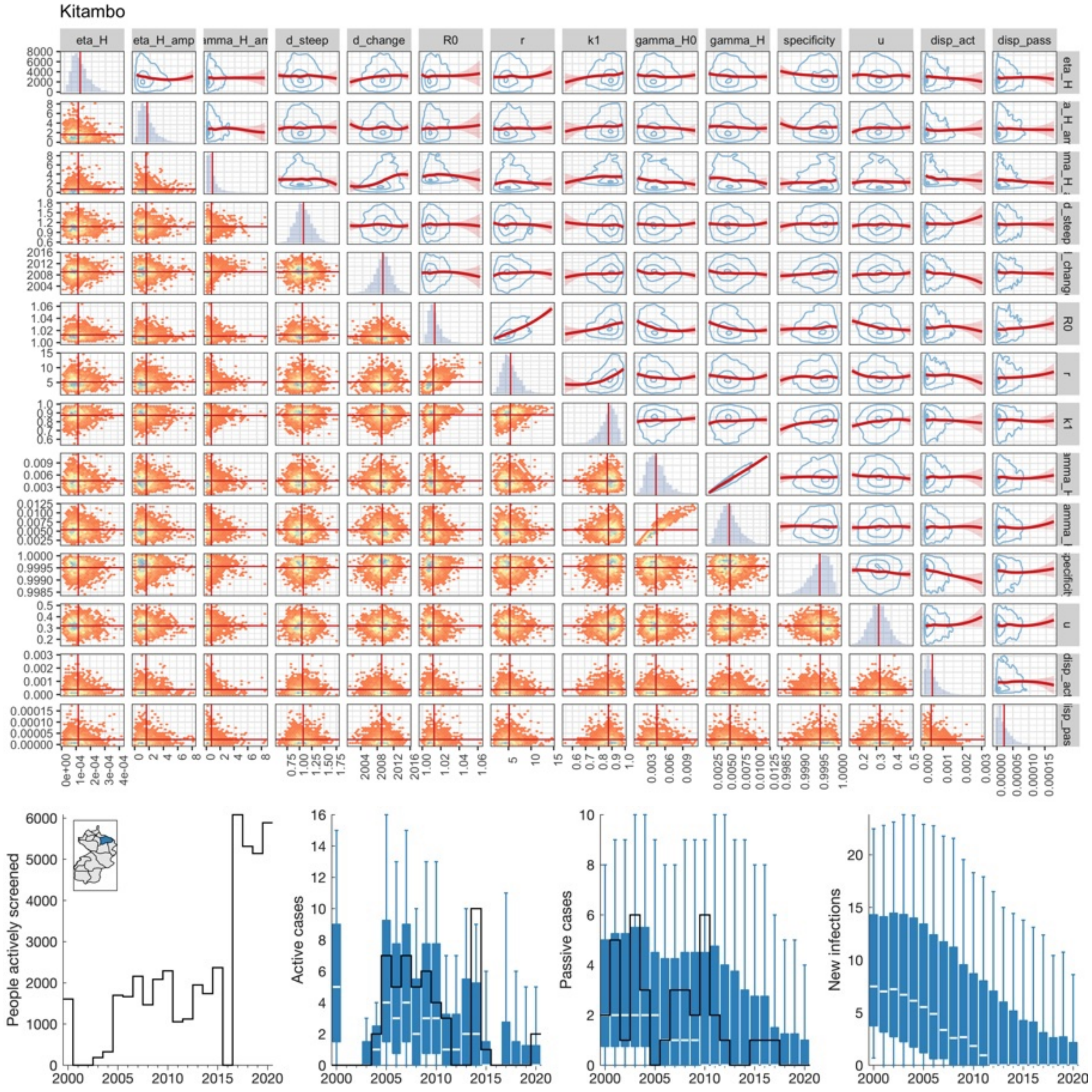


Fig L: **Kitambo (A8) joint posterior and fit to data.**

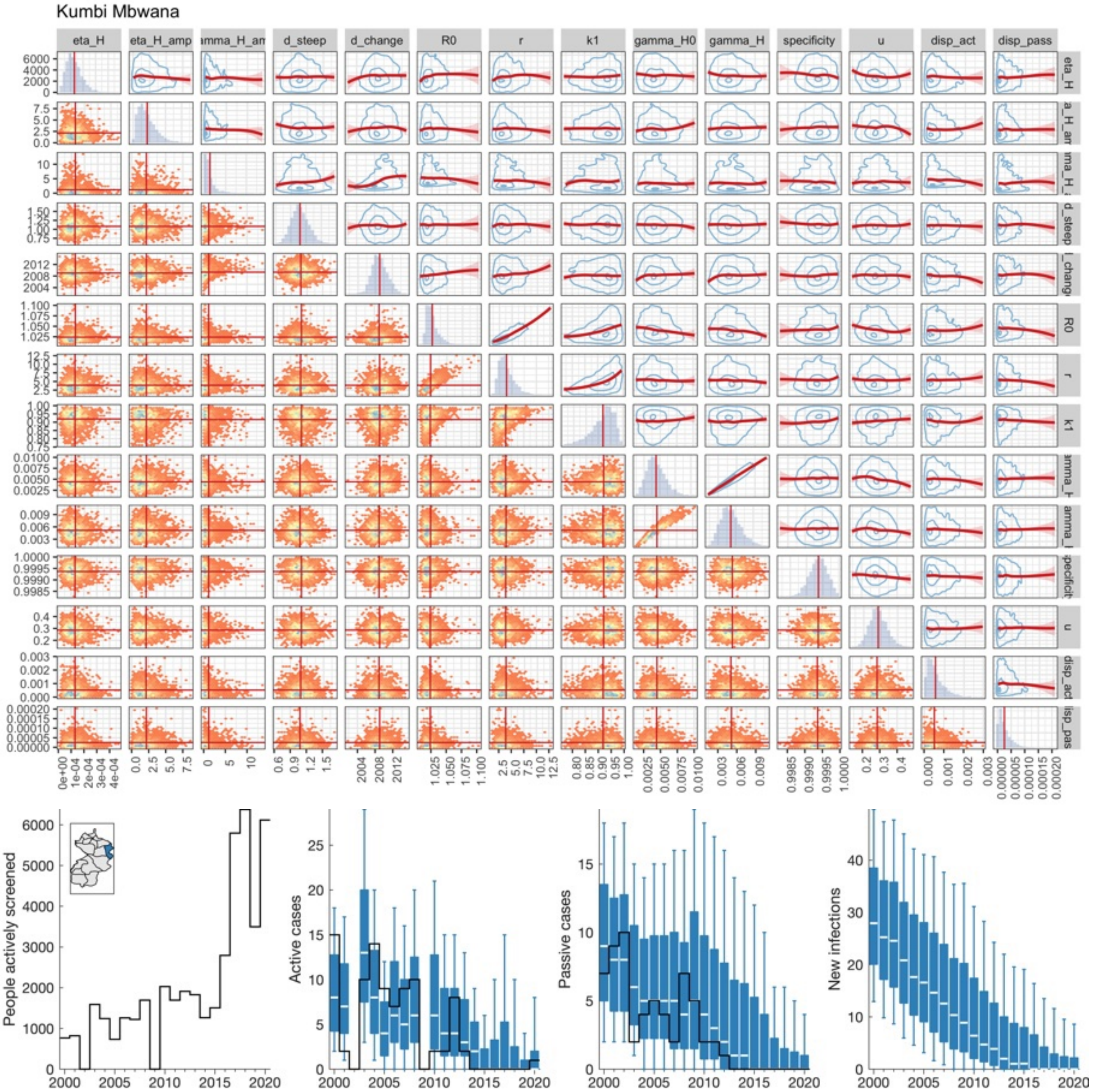


Fig M: **Kumbi Mbwana (A9) joint posterior and fit to data.**

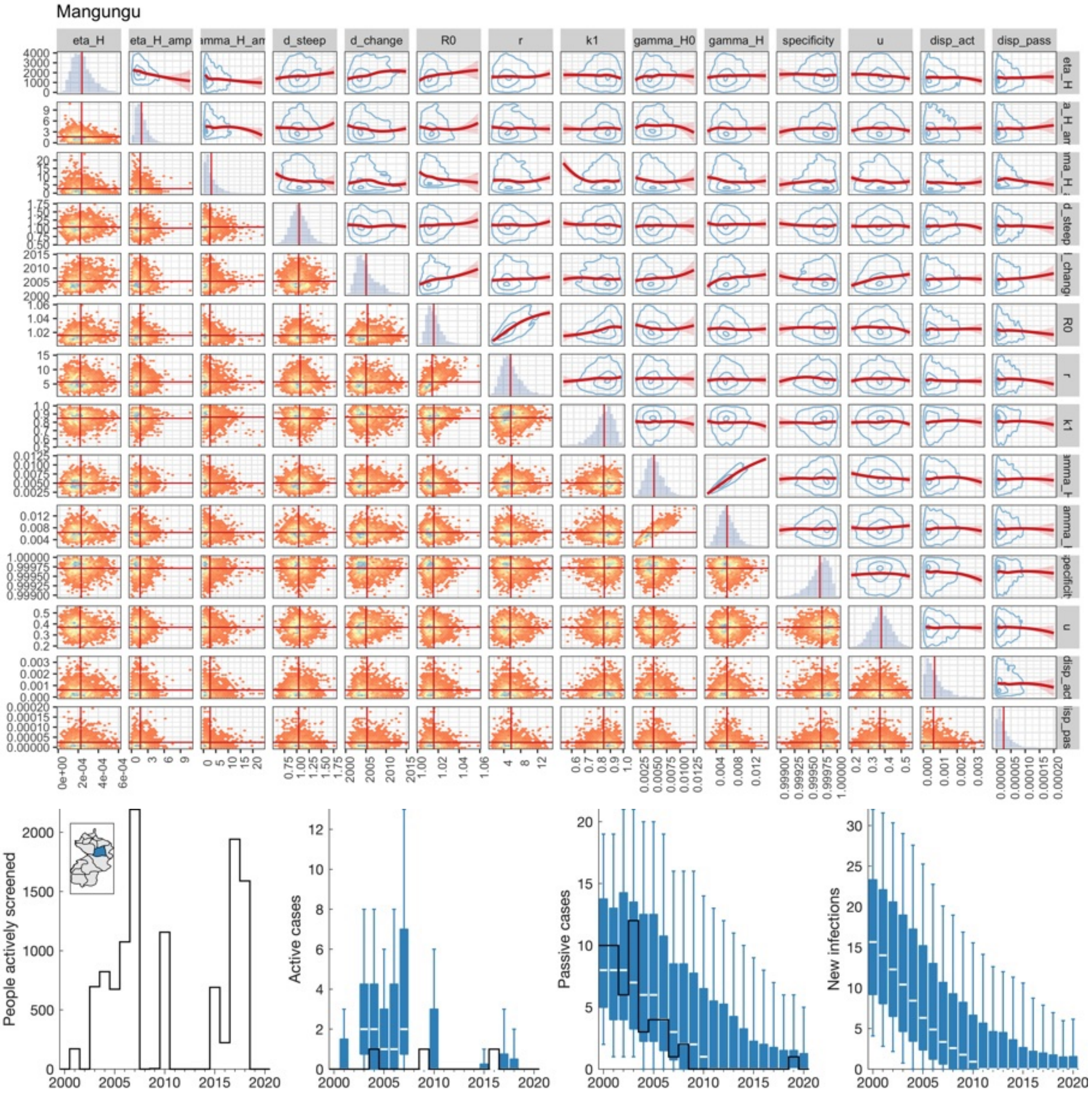


Fig N: **Mangungu (A10) joint posterior and fit to data.**

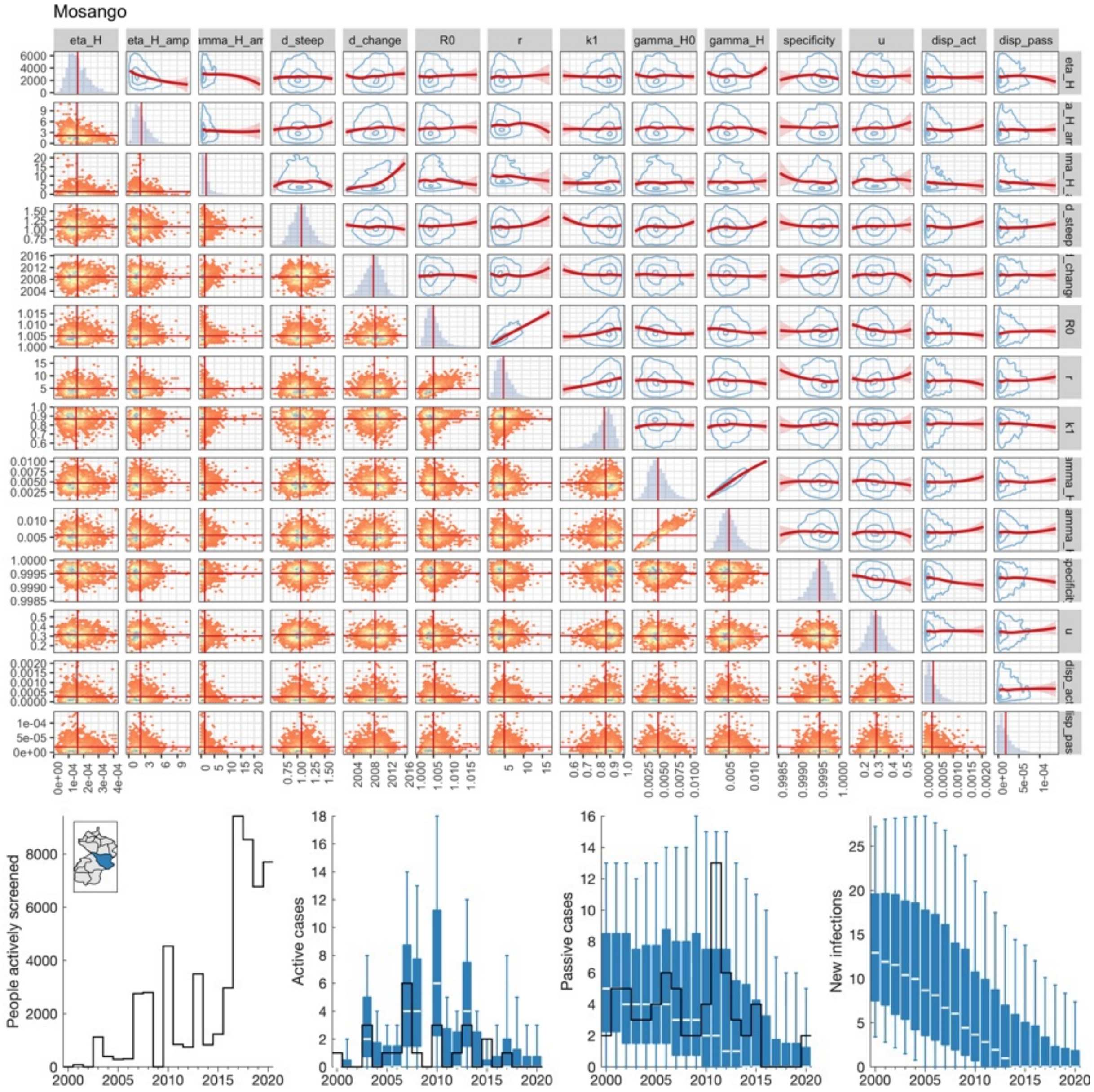


Fig O: **Mosango (A11) joint posterior and fit to data.**

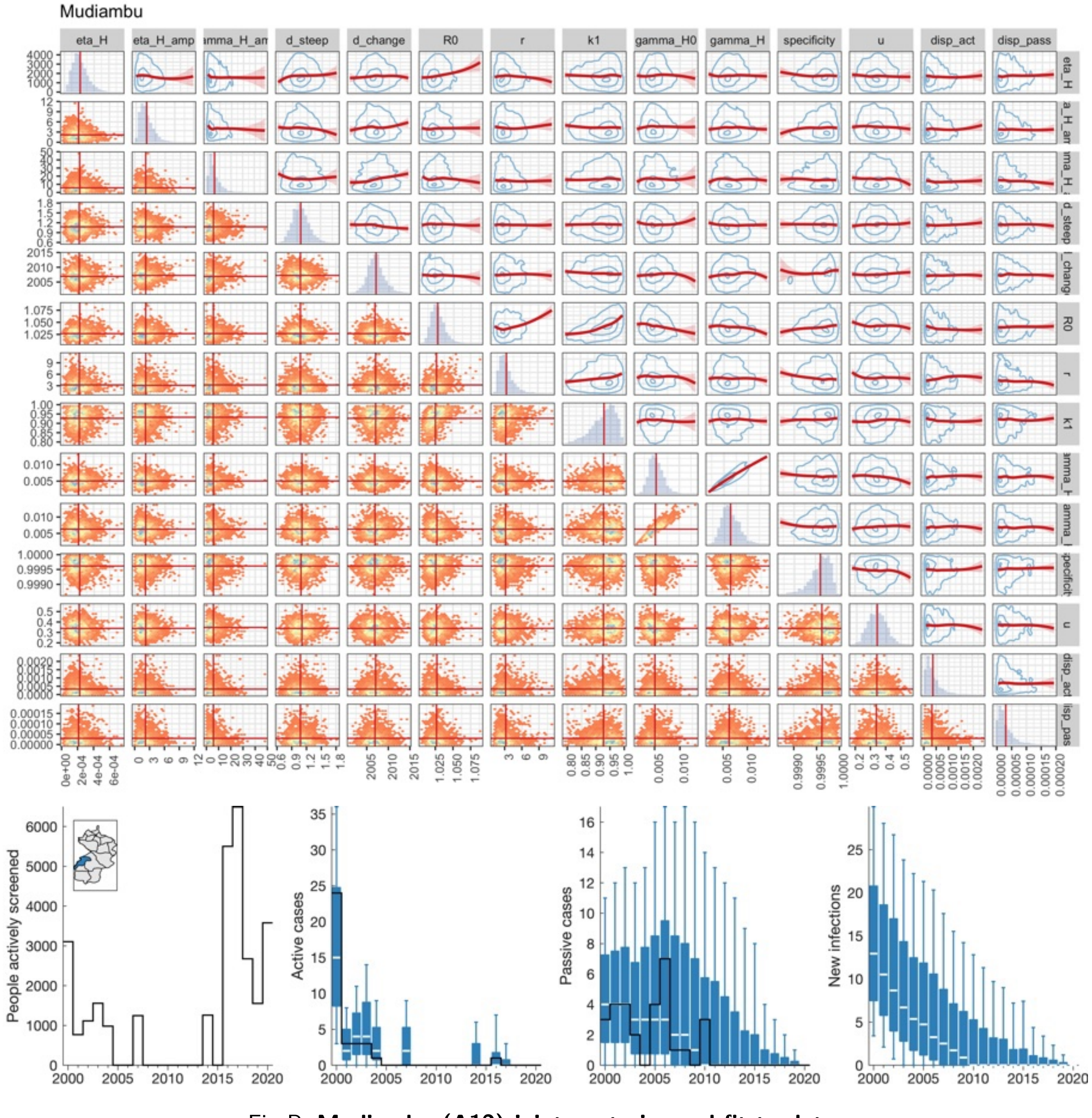


Fig P: **Mudiambu (A12) joint posterior and fit to data.**

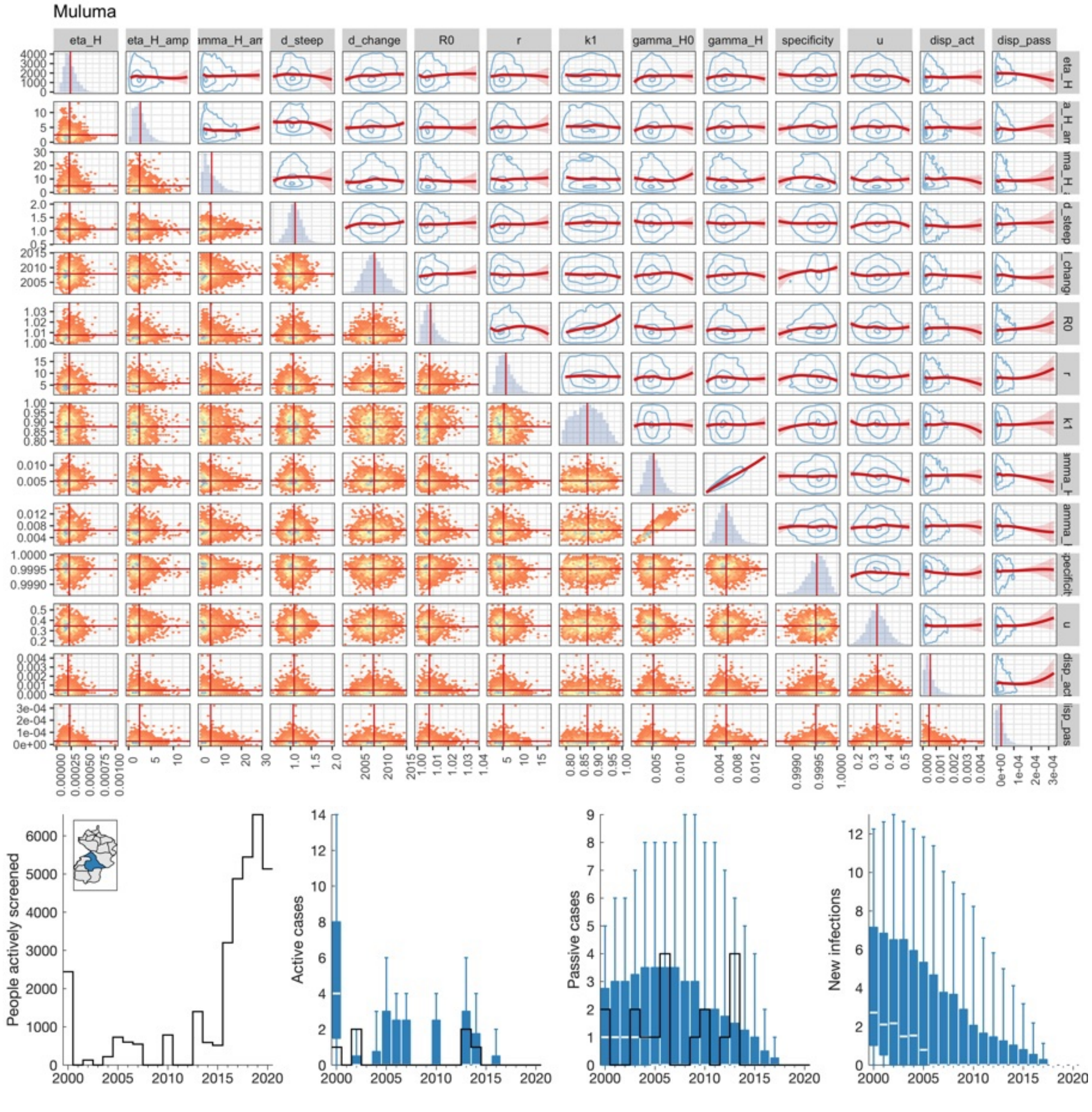


Fig Q: **Muluma (A13) joint posterior and fit to data.**

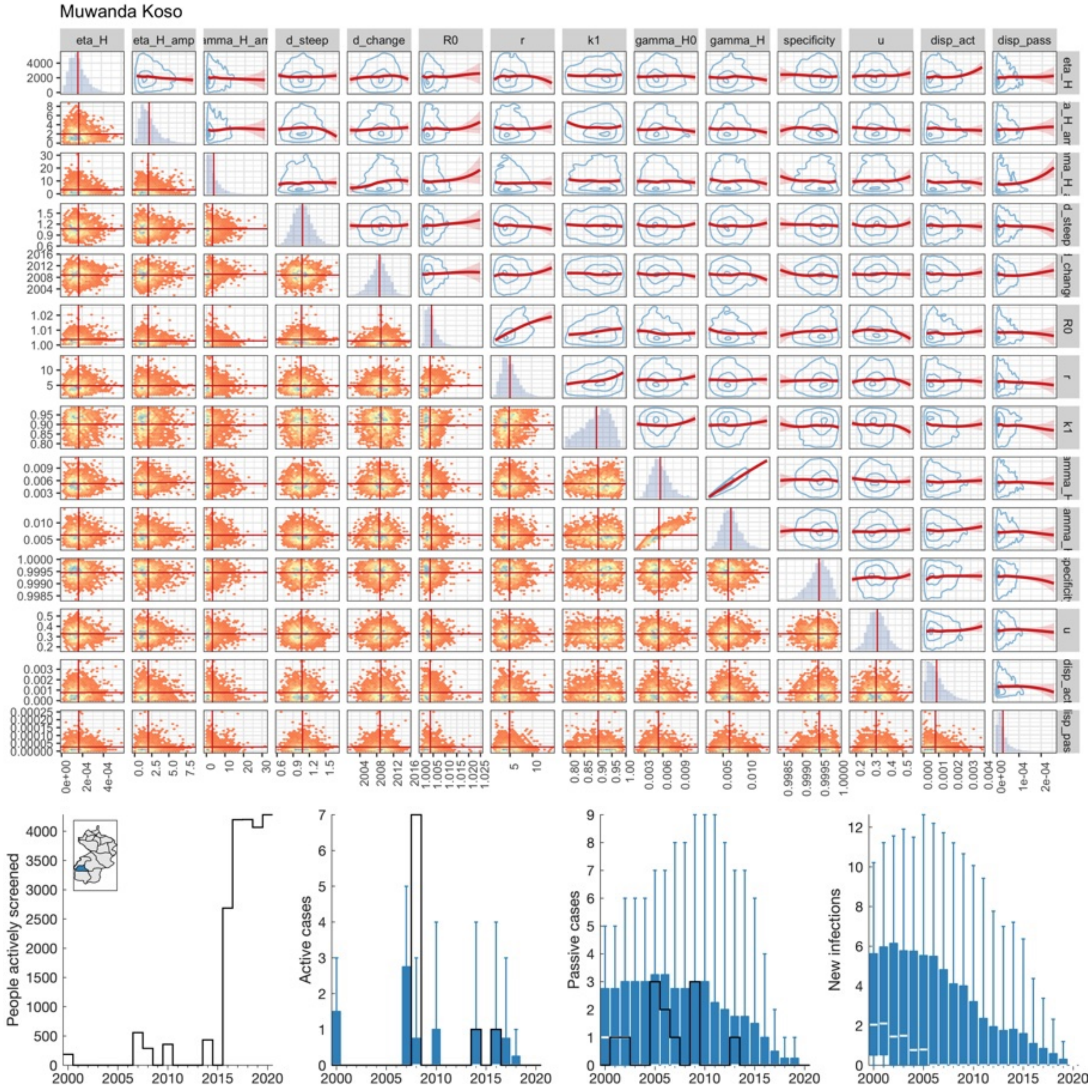


Fig R: **Muwanda Koso (A14) joint posterior and fit to data.**

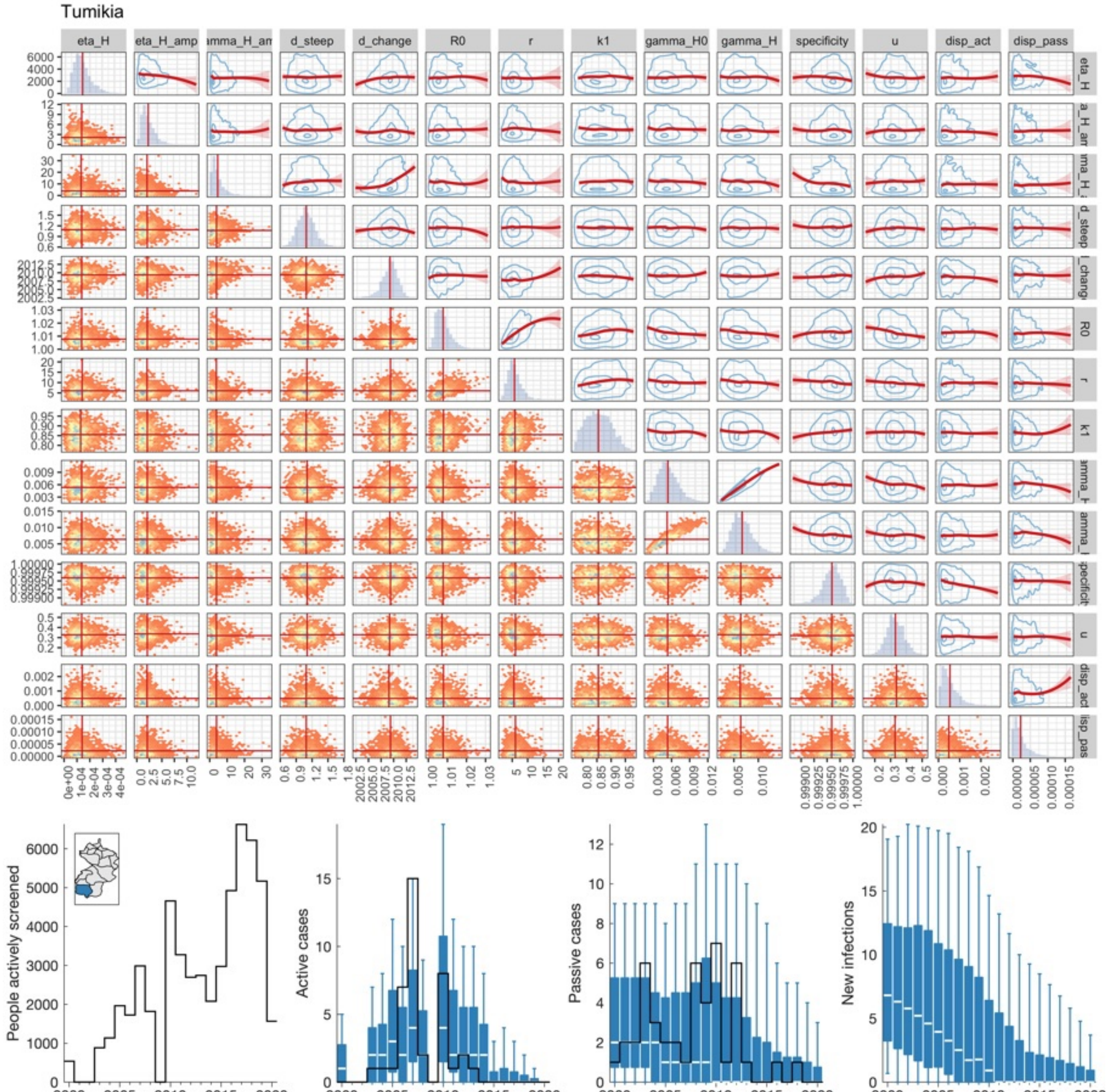


Fig S: **Tumikia (A15) joint posterior and fit to data.**

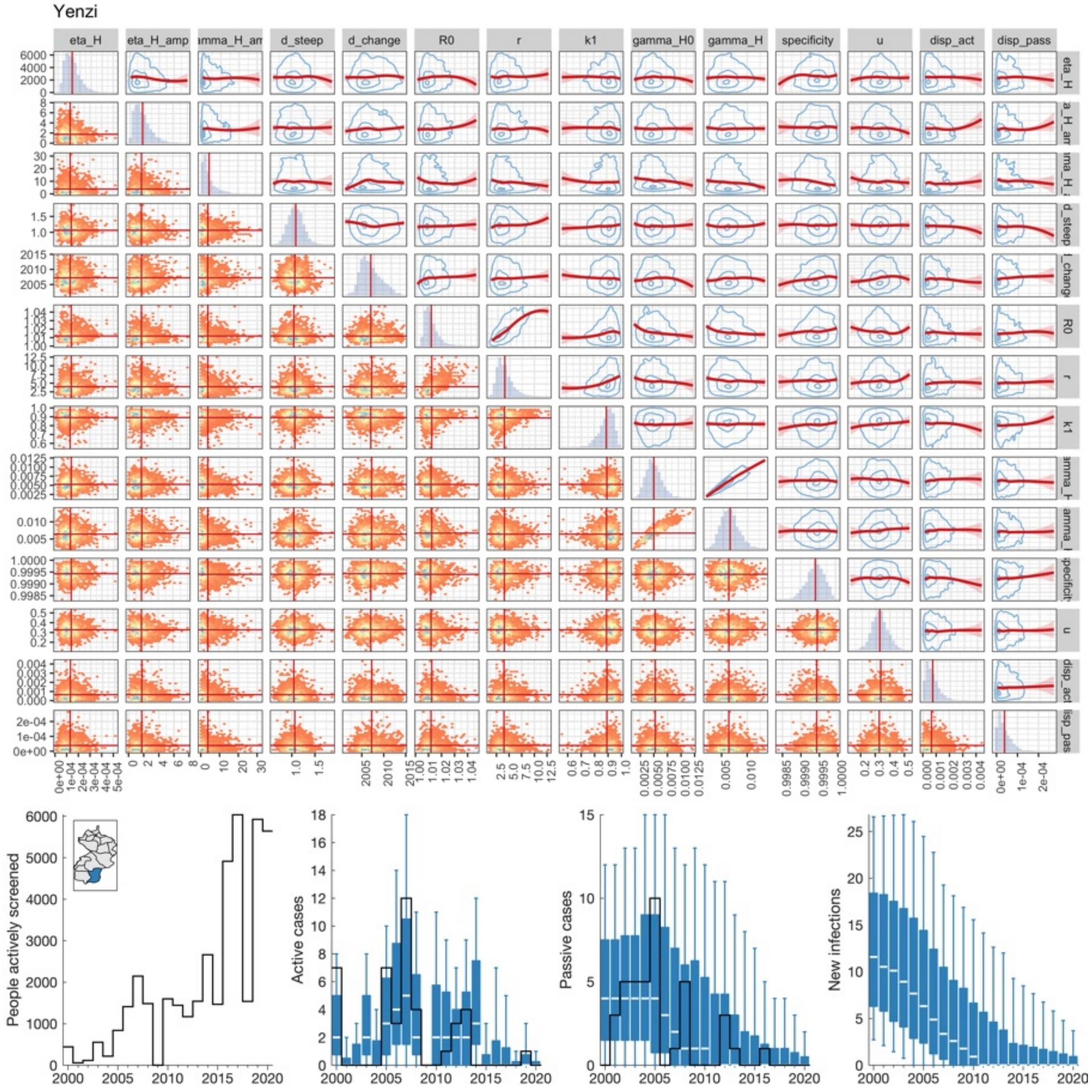


Fig T: **Yenzi (A16) joint posterior and fit to data.**
